# Assessment of Total Parenteral Nutrition administration among Intensive Care Unit patients at Omdurman Military Hospital, Sudan

**DOI:** 10.1101/2020.10.19.20215053

**Authors:** Bothina Essameldin Khalafallah Bashir, Mudawi Mohammed Ahmed Abdallah, Ghada Omer Hamad Abd El-Raheem, Elkhansaa Hamad Ali Nassir

## Abstract

Total parenteral nutrition is one of the important types of nutrition among patients with intestinal failure. This research was intended to assess total parenteral nutrition administration of the intensive care unit patients at the Military hospital, Sudan. A cross-sectional hospital-based study assessed the patients records in the period between April 2014-November 2015, data were analyzed through chi-square test, it was considered significant when *p*≤ 0.05. Twenty patients who received total parenteral nutrition were assessed, 60% were males, while 40% were females. The most frequent indication for total parenteral nutrition was laparotomy (35% of patients). The duration of total parenteral nutrition was assessed, 70% of patients had duration between 1-20 days. Regarding total parenteral nutrition complications, the most frequent complication was hypokalemia (45% of patients), refeeding syndrome occurred in 10 % of patients. A statistically significant association was found between total parenteral nutrition duration when assessed with age and indication (*p*= 0.005 and 0.000 respectively). Patients suffering from electrolytes imbalance need more care to avoid the development of refeeding syndrome, as well as high level of hygiene is strictly required to overcome septic complications. There is a need to consider specialized care team composed of nurses, clinical pharmacists and nutritionists.

## Introduction

Nutritional support can be in the form of oral diet or artificial nutritional support such as enteral feeding or total parenteral nutrition (TPN), TPN is used when the gastro intestinal tract should not be used or do not absorb enough nutrients to maintain adequate nutritional status [1,2]. TPN involves the IV administration of fluids, macronutrients, electrolytes, vitamins and trace elements, for the purpose of weight maintenance or gain, to preserve or restore lean body mass, visceral proteins and to support anabolism [2, 3].

The primary energy source in TPN solutions are carbohydrates, usually as dextrose. Proteins sources are presented as amino acids, in some cases amino acids requirements are higher in patients with burns or undergoing continuous renal replacement therapy [2, 4]. Intravenous fat emulsions (IVFEs) have two main clinical uses, for prevention and treatment of essential fatty acids deficiency, or as a source of energy. IVFE are contraindicated in patients with impaired ability to clear fat emulsions and should be administered cautiously to patients with egg allergy [5]. The micronutrient components of TPN are multivitamins, electrolytes and trace elements [2]. Multiple electrolyte solutions are useful for stable patients with normal organ function who are receiving TPN [6].

TPN additives include heparin, to maintain catheter patency, reduce thrombophlebitis, and to enhance lipid particle clearance [7]. Regular insulin may be added to TPN admixtures for glycemic control, the doses of insulin depend on the severity of hyperglycemia and daily insulin requirements [8, 9, 10].

Refeeding syndrome is one of the complications of TPN administration, it is defined as severe fluid and electrolyte shifts in malnourished patients precipitated by the introduction of nutrition, it may lead to serious disorders such as altered myocardial function, cardiac arrhythmia, hemolytic anemia, liver dysfunction, neuromuscular abnormalities, acute ventilator failure, GT disturbance, renal disorders and even death [11]. Catheter-related blood stream infection (CRBSI) is another TPN complication [12, 13, 14]. Hyperglycemia is common with TPN use, a separate IV insulin infusion is most commonly used for pediatric patients, but it may also provide better and safer glycemic control for patients with very large insulin requirements or unstable marked fluctuations in their blood glucose concentration [15]. Although TPN use is increasing [16], there are few researches done in this scope assessing whether TPN administration was done correctly and whether complications were well managed, this research was intended to assess TPN administration among the intensive care units (ICU) patients of the Military hospital, Sudan.

## Materials and Methods

A hospital-based cross-sectional study assessed the use of TPN among ICU patients based on the medical records of 20 critically ill patients in the intensive care units of the Military Hospital of Khartoum State, Sudan. The Military Hospital is a complex of seven specialized hospitals totalizing 722 beds and 8 ICUs. All 20 adult patients with TPN indication on the period from April 2014 till November 2015 were involved in the study. Data were collected retrospectively, through a standardized questionnaire extracted data from the medical records of ICU patients hospitalized at the time of the data collection. The characteristics of the patients: age, gender, associated comorbidities were recorded. The statistical package for social sciences (SPSS version 23) was used to describe and analyse the data. Statistical analysis test performed was chi-square tests to determine association among variables, it was considered statistically significant when *p* < 0.05.

## Results

### Characteristics of the study participants

20 adult patients receiving TPN were assessed, 40% (8/20) of them aged above 60 years. Males were 60% (12/20), while, females were 40% (8/20). 65% (13/20) of patients weighed between 51-75 Kg. Regarding co-morbidities, 80% (16/20) of patients had no comorbidities, 15% (3/20) were diabetics and 5% (1/20) had CKD. Table 1 below, presents the patients characteristics.

**Table 1:** Characteristics of the study participants (n=20)

| Variable | n | Percent | Variable | n | Percent |
| --- | --- | --- | --- | --- | --- |
| **Age in years:** |  |  | **Co-morbidities:** |  |  |
| 18-30 years | 5 | 25 | Diabetes | 3 | 15 |
| 31-60 years | 7 | 35 | CKD | 1 | 5 |
| > 60 years | 8 | 40 | None | 16 | 80 |
| **Gender:** |  |  |  |  |  |
| Males | 12 | 60 |  |  |  |
| Females | 8 | 40 |  |  |  |
| **Weight in Kg:** |  |  |  |  |  |
| 10-50 Kg | 3 | 15 |  |  |  |
| 51-75 Kg | 13 | 65 |  |  |  |
| 76-100 Kg | 4 | 20 |  |  |  |

\*CKD=Chronic Kidney Disease

### Indications and duration of TPN administration

The indications for TPN use were assessed in our study patients, laparotomy was indicated in 35% (7/20) of cases, and intestinal obstruction was the indication for 30% (6/20) of patients, the remaining other indications are detailed in table 2, below.

**Table 2:** Indications and duration of TPN administration (n=20)

| Variables | n | Percent |
| --- | --- | --- |
| **Indications for TPN** |  |  |
| Laparotomy | 7 | 35 |
| Intestinal obstruction | 6 | 30 |
| Gun shunt in abdomen | 3 | 15 |
| Preparation for surgery | 1 | 5 |
| Refuse oral feeding | 1 | 5 |
| Patient on inotropic agent | 1 | 5 |
| Congenital abnormalities in large bowel | 1 | 5 |
| **Duration of TPN** |  |  |
| 1-20 days | 14 | 70 |
| 21-40 days | 3 | 15 |
| 41-60 days | 2 | 10 |
| 61-80 days | 1 | 5 |

With regards to the duration of TPN administration, 70% (14/20) of patients received TPN for 1 to 20 days, the remaining patients received TPN for longer periods, as detailed in table 2, below.

### Complications of TPN administration

Complications that occurred due to the use of TPN were reported in figure 1 below, hypokalemia was reported in 45% of patients, 40% of patients had hypomagnesemia, hypophosphatemia and line sepsis were noticed in 25% of patients, while refeeding syndrome was reported for 10% of patients. Hyperglycemia was recorded in 15% of patients, all were diabetics, figure 1.

**Figure 1:**
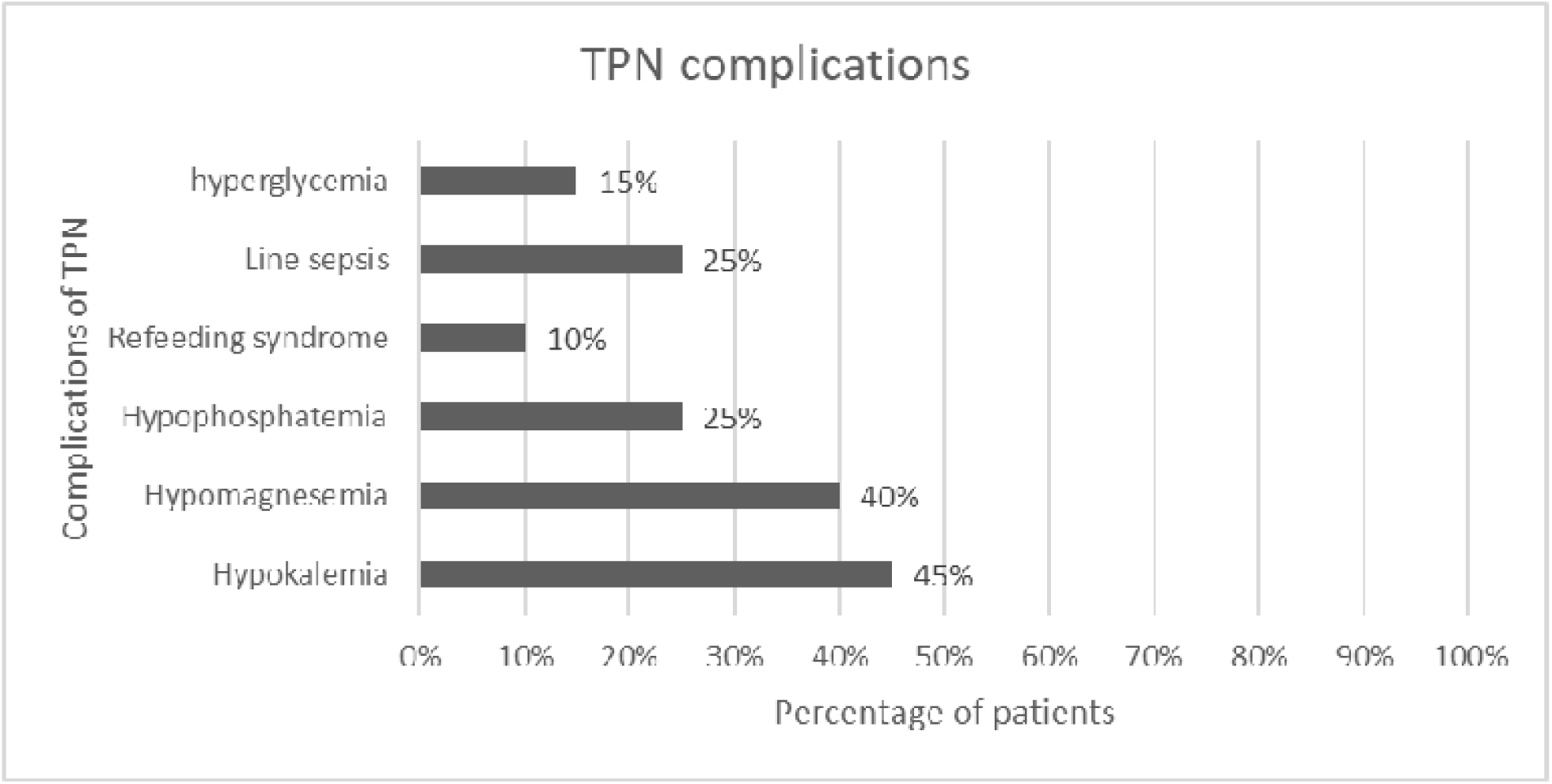
Complications of TPN administration.

### Assessment of the duration of TPN association to the indication and patients characteristics

The association between duration of TPN and age of patients was assessed, a statistically significant association (Likelihood ratio=10.433, *p*-value= 0.005) was found between age and duration of TPN use, table 3. The gender had no statistically significant association with the duration of TPN, as well as, the weight of patients (*p*> 0.05). In the other hand, a statistically significant association (Likelihood ratio=19.936, *p*-value= 0.000) was found between the duration of TPN and the indication, table 3.

**Table 3:** Association between the duration of TPN, indication and patients characteristics (n=20)

| Variables | TPN duration in days |  |  |  | Total | % | Likelihood ratio | p-value |
| --- | --- | --- | --- | --- | --- | --- | --- | --- |
|  | 1-20 d | % | > 20 d | % |  |  |  |  |
| **Age** |  |  |  |  |  |  | 10.433 | 0.005 |
| 18-30 years | 1 | 20.0 | 4 | 80.0 | 5 | 25.0 |  |  |
| 31-60 years | 7 | 100.0 | 0 | 0.0 | 7 | 35.0 |  |  |
| > 60 years | 6 | 75.0 | 2 | 25.0 | 8 | 40.0 |  |  |
| **Total** | **14** | **70.0** | **6** | **30.0** | **20** | **100.0** |  |  |
| **Gender** |  |  |  |  |  |  | 2.106 | 0.147 |
| Male | 7 | 58.3 | 5 | 41.7 | 12 | 60.0 |  |  |
| Female | 7 | 87.5 | 1 | 12.5 | 8 | 40.0 |  |  |
| **Total** | **14** | **70.0** | **6** | **30.0** | **20** | **100.0** |  |  |
| **Weight** |  |  |  |  |  |  | 2.071 | 0.355 |
| 10-50 Kg | 1 | 33.3 | 2 | 66.7 | 3 | 15.0 |  |  |
| 51-75 Kg | 10 | 76.9 | 3 | 23.1 | 13 | 65.0 |  |  |
| 76-100 Kg | 3 | 75.0 | 1 | 25.0 | 4 | 20.0 |  |  |
| **Total** | **14** | **70.0** | **6** | **30.0** | **20** | **100.0** |  |  |
| **TPN Indication** |  |  |  |  |  |  | 19.936 | 0.000 |
| Laparotomy | 7 | 100.0 | 0 | 0.0 | 7 | 35.0 |  |  |
| Intestinal obstruction | 6 | 100.0 | 0 | 0.0 | 6 | 30.0 |  |  |
| Gun shot in abdomen | 0 | 0.0 | 3 | 100.0 | 3 | 15.0 |  |  |
| Others | 1 | 25.0 | 3 | 75.0 | 4 | 20.0 |  |  |
| **Total** | **14** | **70.0** | **6** | **30.0** | **20** | **100.0** |  |  |

## Discussion

Forty percent of our study participants were older patients, aged above 60 years, this was consistent with studies [11, 17, 18, 19]. Males who received TPN were more than females, similar to [11, 19] and controversial to another study [17]. 65% of our study population had normal body weight, the ones with reduced body weight were 15%, a Korean study reported that the body weight ranged between 52-82 Kg [18]. Majority of the participants (80%) had no co-morbidities and only 5% had CKD, in contrast to other study in which CKD was prevalent in 17% of patients [17]. In another Chinese study, patients with CKD were 4.8% and 19.6% in the two TPN groups studied [19].

Regarding the duration of TPN, 70% of our patients received TPN for 20 days or less, in other studies, TPN duration was 15 days and 10 days [11, 17]. On assessment of the duration of TPN treatment, with regards to age, majority of patients who received prolonged durations of TPN (> 20 days) were young aged between 18 to 30 years (*p*= 0.005), but when compared to gender, there was no difference in the duration of TPN between males and females (*p*> 0.05).

On assessing TPN indications, laparotomy and intestinal obstruction were the most frequent indications for TPN (35%, 30% respectively), in other study, GI surgeries were more than half of the indications and intestinal obstruction represented only 19% [17]. In our study, these common indications had lesser duration of TPN treatment than other indications such as gunshot wounds in the abdomen which required longer durations of TPN (*p*= 0.000).

Complications of TPN administration, mentioned in published literature [11, 18] were present in our patients, the most frequent complications were the electrolyte imbalances with hypokalemia being the most frequent followed by hypomagnesemia then hypophosphatemia, these outcomes were similar to the published cohort study [11]. In our study, line sepsis was reported in 25% of patients, in another study it was reported in 35.7% and 17.4% of TPN patients [19], and because of the importance of this issue, the causing organisms were studied [20]. This develops concerns about the applications of aseptic techniques by the staff, which might necessitate recruitment of specialized nursing staff to monitor and prevent these septic events [14]. Hyperglycemia was reported in 15% of our study patients and all were diabetics, while in other study, hyperglycemia was reported for 29.6% of patients and 16% of them were diabetics [17]. This seeks needs to have individualized hyperglycemia control protocols for such special populations as in published studies [8, 9,10].

Not to forget, refeeding syndrome was also common among our study population, 10% of patients suffered from refeeding syndrome, this was five times higher than what was published in a cohort study [11], this issue was also tackled in published literature [2, 3, 21]. These complications derives attention to consider TPN care team involving nutrition personnel [22], specialized nursing [14], as well as, clinical pharmacists after assessing all the challenges, similarly to a published study in Kuwait [23].

All this intensifies the need for guidance to the working staff to follow specialized guidelines as A.S.P.E.N and SCCM [1, 24].

The main limitation in our study is the retrospective nature of data collection from medical records, this retrospective method highly relay’s on medical documentations which might not be enough in all cases. Other limitation includes being conducted in a single center.

## Conclusions

Most common indications for TPN administration were laparotomy and intestinal obstruction. Diabetic patients have high risk of developing hyperglycemia. Refeeding syndrome an line sepsis were common among the ICU patients, which needs special consideration such as education of staff, assignment of TPN care team.

## Recommendations

TPN administration need to be confirmed according to the patients need only. Diabetic patients need hyperglycemia control during TPN administration through continuous infusion of insulin.

Patients suffering from electrolytes imbalance need more care to avoid the development of refeeding syndrome. Enrollment of clinical pharmacists to control the administration of TPN is needed. As well as, high level of hygiene is, strictly, required to overcome these complications.

## Data Availability

all data are available in the manuscript

## Data Availability

all data are available in the manuscript

## Declarations

### Ethical approval and consent to participate

Institutional Review Board of Ahfad university for women reviewed the proposal. Ethical approval was obtained from the Military Hospital, the implementation of the research was granted by the administration of the respective ICUs. Regarding participants, their confidentiality was assured with the use of an anonymous research tool and the collected data would be used strictly for the purpose of the study objectives.

### Competing interests

The authors declared no competing interest.

### Funding

Bothina Essameldin Khalafallah Bashir bore all the costs related to the study.

### Authors’ contributions

**BEKB**: Elaborated the research proposal, implemented the field data collection and participated in the manuscript writing.

**AMAM**: Facilitated the administrative arrangements at the Military Hospital, co-supervised and revised the proposal and read the final manuscript prior to submission.

**AOHG**: Conducted the statistical analysis and drafted the initial manuscript and the final manuscript.

**NAHE**: Supervised the implementation of the research from proposal to completion.

## Abbreviations

ASPEN: American society for parenteral and enteral nutrition
CRBSI: Catheter Related Blood Stream Infection
CKD: Chronic kidney disease
ICU: Intensive Care Unit
IV: Intra-Venous
IVFEs: Intra-Venous Fat Emulsions
SCCM: Society of critical care medicine
TPN: Total Parenteral Nutrition

## Acknowledgments

The authors are grateful to the patients of the Military Hospital whose participation enabled this study to be completed.

## Notes

### Competing Interest Statement

The authors have declared no competing interest.

